# Analytical sensitivity and efficiency comparisons of SARS-COV-2 qRT-PCR primer-probe sets

**DOI:** 10.1101/2020.03.30.20048108

**Authors:** Chantal B.F. Vogels, Anderson F. Brito, Anne L. Wyllie, Joseph R. Fauver, Isabel M. Ott, Chaney C. Kalinich, Mary E. Petrone, Arnau Casanovas-Massana, M. Catherine Muenker, Adam J. Moore, Jonathan Klein, Peiwen Lu, Alice Lu-Culligan, Xiaodong Jiang, Daniel J. Kim, Eriko Kudo, Tianyang Mao, Miyu Moriyama, Ji Eun Oh, Annsea Park, Julio Silva, Eric Song, Takehiro Takahashi, Manabu Taura, Maria Tokuyama, Arvind Venkataraman, Orr-El Weizman, Patrick Wong, Yexin Yang, Nagarjuna R. Cheemarla, Elizabeth B. White, Sarah Lapidus, Rebecca Earnest, Bertie Geng, Pavithra Vijayakumar, Camila Odio, John Fournier, Santos Bermejo, Shelli Farhadian, Charles S. Dela Cruz, Akiko Iwasaki, Albert I. Ko, Marie L. Landry, Ellen F. Foxman, Nathan D. Grubaugh

**Author notes:** Correspondence (CBFV); (NDG).

## Abstract

The recent spread of severe acute respiratory syndrome coronavirus 2 (SARS-CoV-2) exemplifies the critical need for accurate and rapid diagnostic assays to prompt clinical and public health interventions. Currently, several quantitative reverse-transcription polymerase chain reaction (qRT-PCR) assays are being used by clinical, research, and public health laboratories. However, it is currently unclear if results from different tests are comparable. Our goal was to evaluate the primer-probe sets used in four common diagnostic assays available on the World Health Organization (WHO) website. To facilitate this effort, we generated RNA transcripts to be used as assay standards and distributed them to other laboratories for internal validation. We then used (1) RNA transcript standards, (2) full-length SARS-CoV-2 RNA, (3) pre-COVID-19 nasopharyngeal swabs, and (4) clinical samples from COVID-19 patients to determine analytical efficiency and sensitivity of the qRT-PCR primer-probe sets. We show that all primer-probe sets can be used to detect SARS-CoV-2 at 500 virus copies per reaction, except for the RdRp-SARSr (Charité) confirmatory primer-probe set which has low sensitivity. Our findings characterize the limitations of currently used primer-probe sets and can assist other laboratories in selecting appropriate assays for the detection of SARS-CoV-2.

## Introduction

Accurate diagnostic assays and large-scale testing are critical for mitigating outbreaks of infectious diseases. Early detection prompts public health actions to prevent and control the spread of pathogens. This has been exemplified by the novel coronavirus, known as SARS-CoV-2, which was first identified as the cause of an outbreak of pneumonia in Wuhan, China, in December 2019, and rapidly spread around the world^1–3^. The first SARS-CoV-2 genome sequence was critical for the development of diagnostics^2^, which led to several molecular assays being developed to detect COVID-19 cases^4–7^. The World Health Organization (WHO) currently lists seven molecular assays (i.e. qRT-PCR) to diagnose COVID-19^8^; however, it is not clear to many laboratories or public health agencies which assay they should adopt.

Our goal was to compare the analytical efficiencies and sensitivities of the primer-probe sets used in the four most common SARS-CoV-2 qRT-PCR assays developed by the China Center for Disease Control (China CDC)^7^, United States CDC (US CDC)^6^, Charité Institute of Virology, Universitätsmedizin Berlin (Charité)^5^, and Hong Kong University (HKU)^4^. To this end, we first generated RNA transcripts from a SARS-CoV-2 isolate from an early COVID-19 case from the state of Washington (United States)^9^. Using RNA transcripts, SARS-CoV-2 RNA from cell culture, pre-COVID-19 nasopharyngeal swabs, and clinical samples from COVID-19 patients, we find mostly similar analytical sensitivity of the tested primer-probe sets to detect low amounts of SARS-CoV-2, with the exception of the Charité confirmatory assay. Thus, we provide insights in analytical efficiency and sensitivity of commonly used primer-probe sets that should be considered when selecting and validating appropriate assays for SARS-CoV-2 detection.

## Results

### Generation of RNA transcript standards for qRT-PCR validation

A barrier to implementing and validating qRT-PCR molecular assays for SARS-CoV-2 detection was the availability of virus RNA standards. As the full length SARS-CoV-2 RNA is considered as a biological safety level 2 hazard in the US, we generated small RNA transcripts (704-1363 nt) from the non-structural protein 10 (nsp10), RNA-dependent RNA polymerase (RdRp), non-structural protein 14 (nsp14), envelope (E), and nucleocapsid (N) genes spanning each of the primer and probe sets in the China CDC^7^, US CDC^6^, Charité^5^, and HKU^4^ assays (**Fig. 1A**; **Table 1; Supplemental Tables 1-2**)^10^. By measuring PCR amplification using 10-fold serial dilutions of our RNA transcript standards, we found the efficiencies of each of the nine primer-probe sets to be above 90% (**Fig. 1B**), which match the criteria for an efficient qRT-PCR assay^11^. Our RNA transcripts can thus be used for assay validation, positive controls, and standards to quantify viral loads: critical steps for a diagnostic assay. Our protocol to generate the RNA transcripts is openly available^10^, and any clinical or research diagnostic lab can directly request them for free through our lab website (www.grubaughlab.com).

**Table 1:**
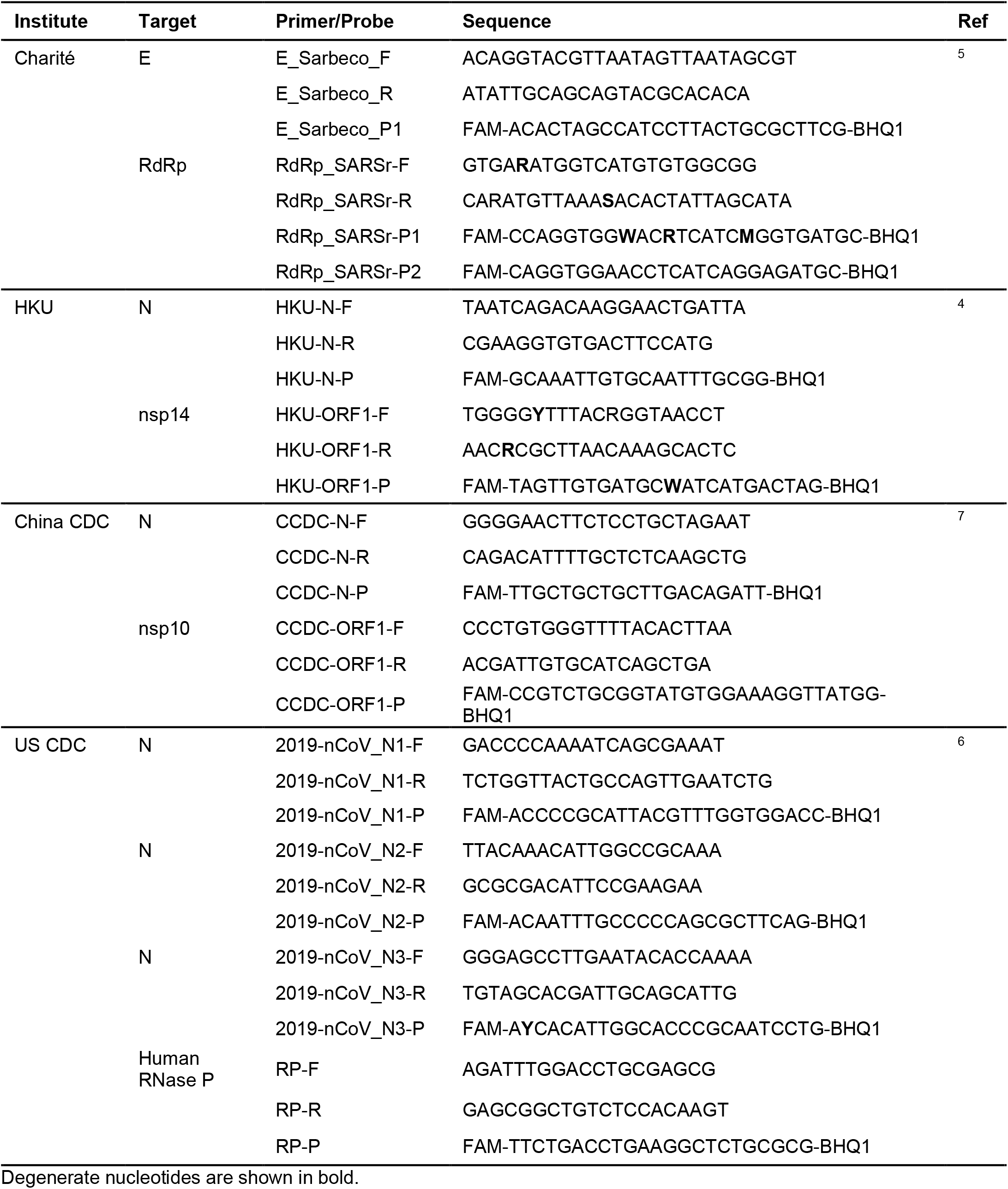
Primers and probes for common SARS-CoV-2 qRT-PCR diagnostic assays.

**Fig. 1:**
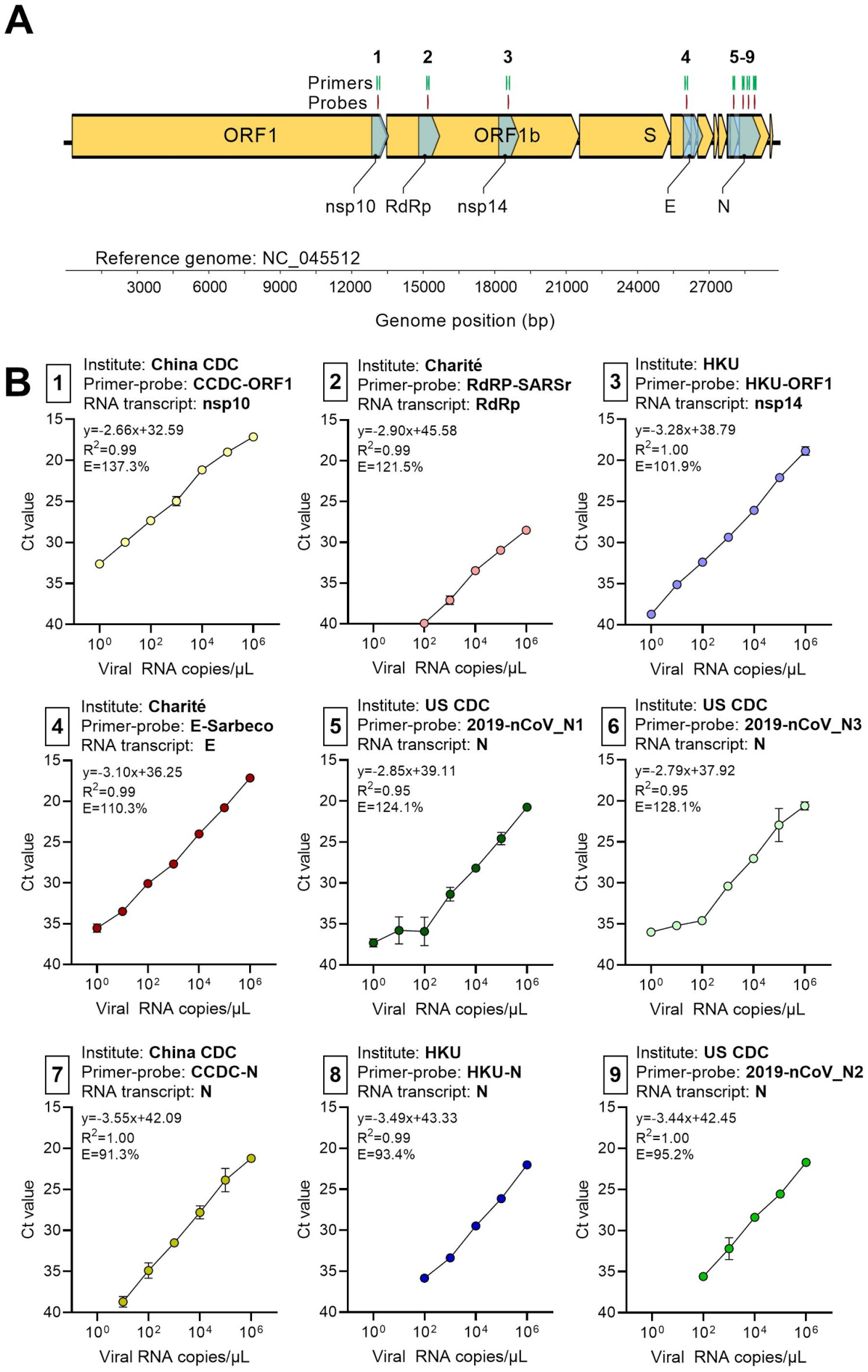
Generation of RNA transcript standards for validation of SARS-CoV-2 qRT-PCR assays. (**A**) We reverse-transcribed RNA transcript standards for the non-structural protein 10 (nsp10), RNA-dependent RNA polymerase (RdRp), non-structural protein 14 (nsp14), envelope (E), and nucleocapsid (N) genes for validation of nine primer-probe sets used in SARS-CoV-2 qRT-PCR assays. (**B**) We generated standard curves for nine primer-probe sets with 10-fold dilutions (10^0^-10^6^ viral RNA copies/μL) of the corresponding RNA transcript standards. For each combination of primer-probe set and RNA transcript standard, we provide the slope, intercept, R^2^, and efficiency. Error bars show the standard deviation. The primer-probe sets are numbered as shown in panel A.

### Analytical comparisons of qRT-PCR primer-probe sets

Critical evaluations of the designed primer-probe sets used in the primary SARS-CoV-2 qRT-PCR detection assays are necessary to compare findings across studies, and select appropriate assays for in-house testing. The goal of our study was to compare the designed primer-probe sets, not the assays *per se*, as that would involve many different variables. To do so we used the same (***1***) primer-probe concentrations (500 nM of forward and reverse primer, and 250 nM of probe); (***2***) PCR reagents (New England Biolabs Luna Universal Probe One-step RT-qPCR kit); and (***3***) thermocycler conditions (40 cycles (45 for clinical samples) of 10 seconds at 95°C and 20 seconds at 55°C) in all reactions. From our measured PCR amplification efficiencies and analytical sensitivities of detection, most primer-probe sets were comparable, except for the RdRp-SARSr (Charité) set, which had low sensitivity (**Fig. 2**).

**Fig. 2:**
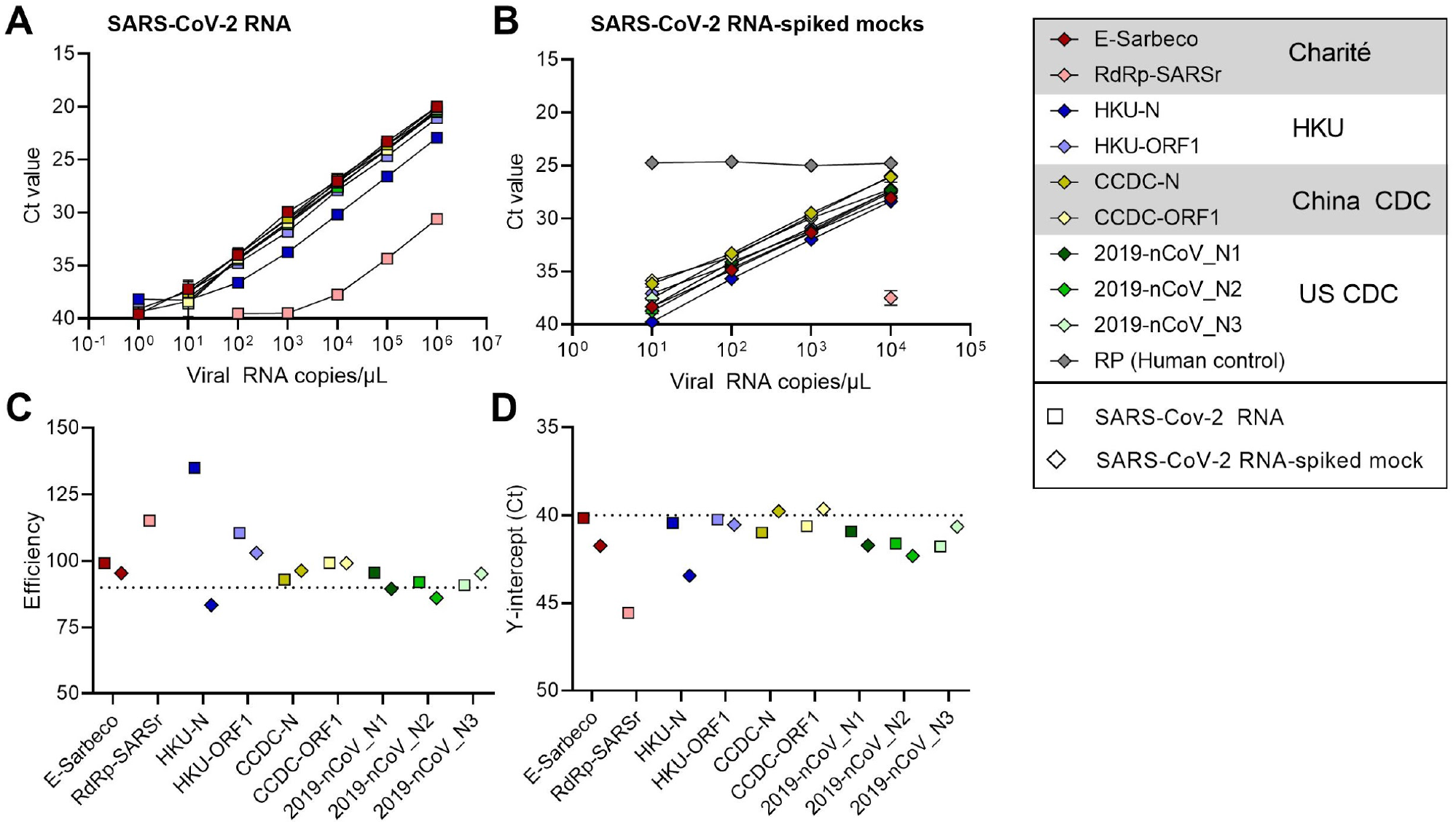
Analytical efficiency and sensitivity of the nine primer-probe sets used in SARS-CoV-2 qRT-PCR assays. We compared nine primer-probe sets and a human control primer-probe set targeting the human RNase P gene with 10-fold dilutions of (**A**) full-length SARS-CoV-2 RNA and (**B**) pre-COVID-19 mock samples spiked with known concentrations of SARS-CoV-2 RNA. We determined (**C**) efficiency and (**D**) y-intercept Ct values (measured analytical sensitivity) of the nine primer-probe sets. We extracted nucleic acid from SARS-CoV-2-negative nasopharyngeal swabs (collected from respiratory disease patients in 2017) and spiked these with known concentrations of SARS-CoV-2 RNA. Symbols depict sample types: squares represent tests with SARS-CoV-2 RNA and diamonds represent SARS-CoV-2 RNA-spiked mock samples. Colors depict the nine tested primer-probe sets. Error bars show the standard deviation. The CDC human RNase P (RP) assay was included as an extraction control.

By testing each of the nine primer-probe sets using 10-fold dilutions of SARS-CoV-2 RNA derived from cell culture (**Fig. 2A**) or 10-fold dilutions of SARS-CoV-2 RNA spiked into RNA extracted from pooled nasopharyngeal swabs taken from patients in 2017 (SARS-CoV-2 RNA-spiked mocks; **Fig. 2B**), we again found that the PCR amplification efficiencies were near or above 90% (**Fig. 2C**). To measure the analytical sensitivity of virus detection, we used the cycle threshold (Ct) value in which the expected linear dilution series would cross the y-intercept when tested with 1 viral RNA copy per μL of RNA. Our measured sensitivities (y-intercept Ct values) were similar among most of the primer-probe sets, except for the RdRp-SARSr (Charité) set (**Fig. 2D**). We found that the Ct values from the RdRp-SARSr set were usually 6-10 Cts higher (lower virus detection) than the other primer-probe sets.

### Detection of virus at low concentrations

To determine the lower limit of detection and the occurrence of false positive or inconclusive detections, we tested the primer-probe sets using SARS-CoV-2 RNA spiked into RNA extracted from pooled nasopharyngeal swabs from respiratory disease patients during 2017 (pre-COVID-19). Our mock clinical samples demonstrated that all primer-probe sets, except RdRp-SARSr (Charité), had a lower detection limit of 100 viral RNA copies per μL (500 copies/reaction; **Fig. 3**).

**Fig. 3:**
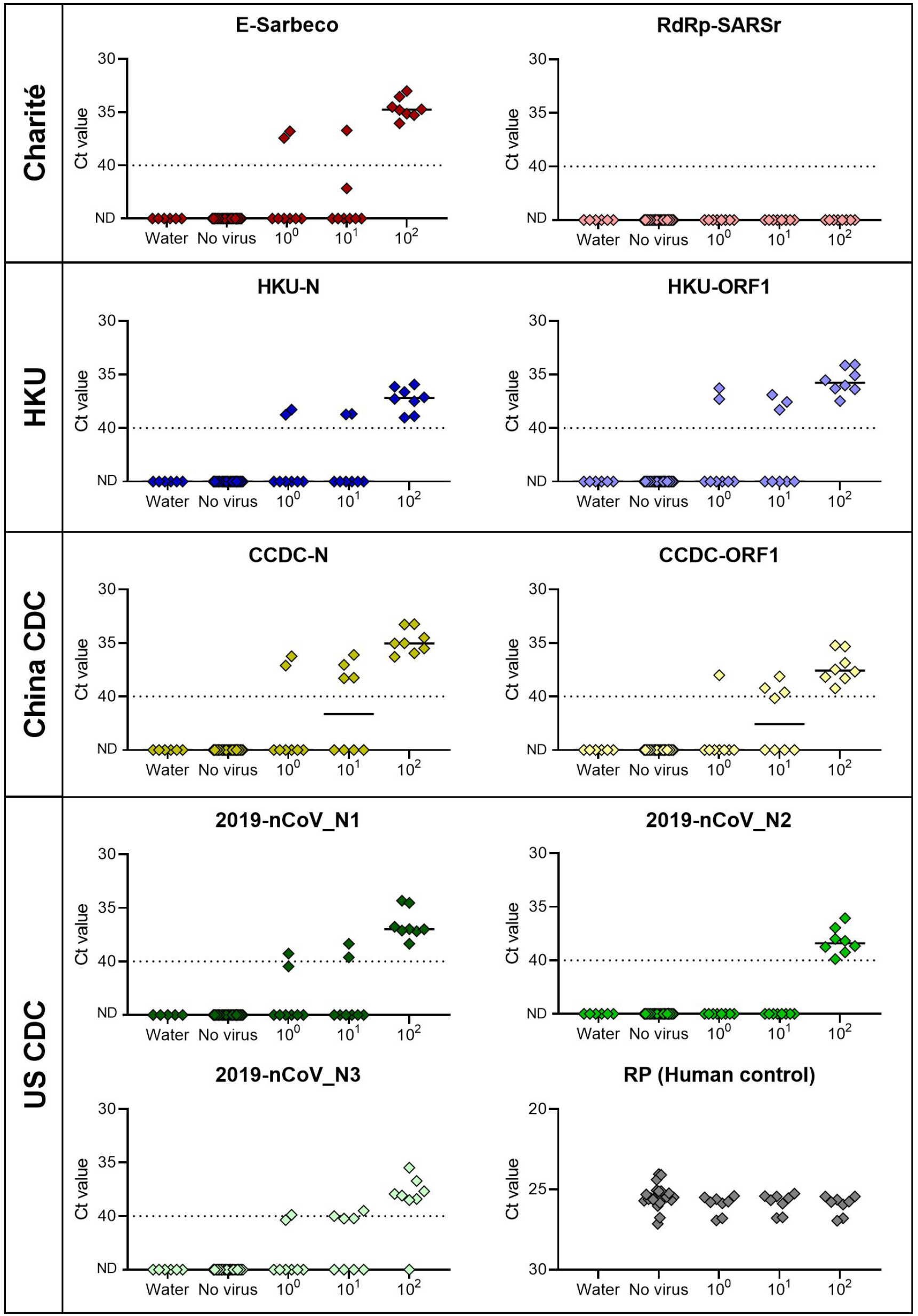
All primer-probe sets, except RdRp-SARSr, have a lower detection limit of 10^2^ SARS-CoV-2 viral RNA copies/μL spiked into pre-COVID-19 nasopharyngeal swabs. We determined the lower detection limit of nine primer-probe sets as well as the human RNase P control for mock samples (RNA extracted from nasopharyngeal swabs collected in 2017) spiked with known concentrations of SARS-CoV-2 RNA. We performed 24 technical replicates with mock samples without spiking RNA (no virus) and 8 replicates (2 replicates with 4 independent pools of each 4 nasopharyngeal swabs) with mock samples spiked with 10^0^-10^2^ viral RNA copies/μL of SARS-CoV-2 RNA. ND = not detected. Black lines indicate the median and the dashed line indicates the detection limit.

When testing nasopharyngeal swabs collected prior to the COVID-19 pandemic, we found that qRT-PCR did not result in background amplification for any of the tested primer-probe sets (**Fig. 3**). These findings suggest that there is no cross-reactivity between the tested primer-probe sets and possible host or pathogen nucleic acid present in nasopharyngeal swabs collected pre-COVID-19. When using SARS-CoV-2 RNA spiked into RNA from these nasopharyngeal swabs, our results show that all primer-probe sets, except RdRp-SARSr and 2019-nCoV_N2, were able to partially detect (Ct values <40) SARS-CoV-2 RNA at 1 (10^0^) and 10 (10^1^) viral RNA copies/μL (**Fig. 3**). Thus, our results show that there is very limited variation between analytical sensitivity of the tested primer-probe sets. At 100 (10^2^) viral RNA copies/μL, we could detect virus and differentiate between the negative samples for all primer-probe sets, except for the RdRp-SARSr (Charité) set, which was negative (Ct values >40) for all 10^0^-10^2^ viral RNA copies/μL concentrations.

### Performance of US CDC primer-probe sets with clinical samples

We found slight differences in analytical sensitivity of the 2019-nCoV_N1 and N2 primer-probe sets that currently comprise the US CDC assay. We were able to partially detect SARS-CoV-2 RNA spiked into 2017 nasopharyngeal swabs up to 1 viral RNA copy/μL with N1 whereas we didn’t detect positives below 100 viral RNA copies/μL for N2. To investigate the potential impact of small differences in analytical sensitivity between N1 and N2 on test outcomes, we compared 2019-nCoV_N1 and N2 results from 172 clinical samples taken during the COVID-19 pandemic (**Fig. 4**). We found that N1 was typically more sensitive, yielding lower Ct values from positive samples, but that the combination of both primer-probe sets did not yield an abundance of inconclusive test results.

**Fig. 4:**
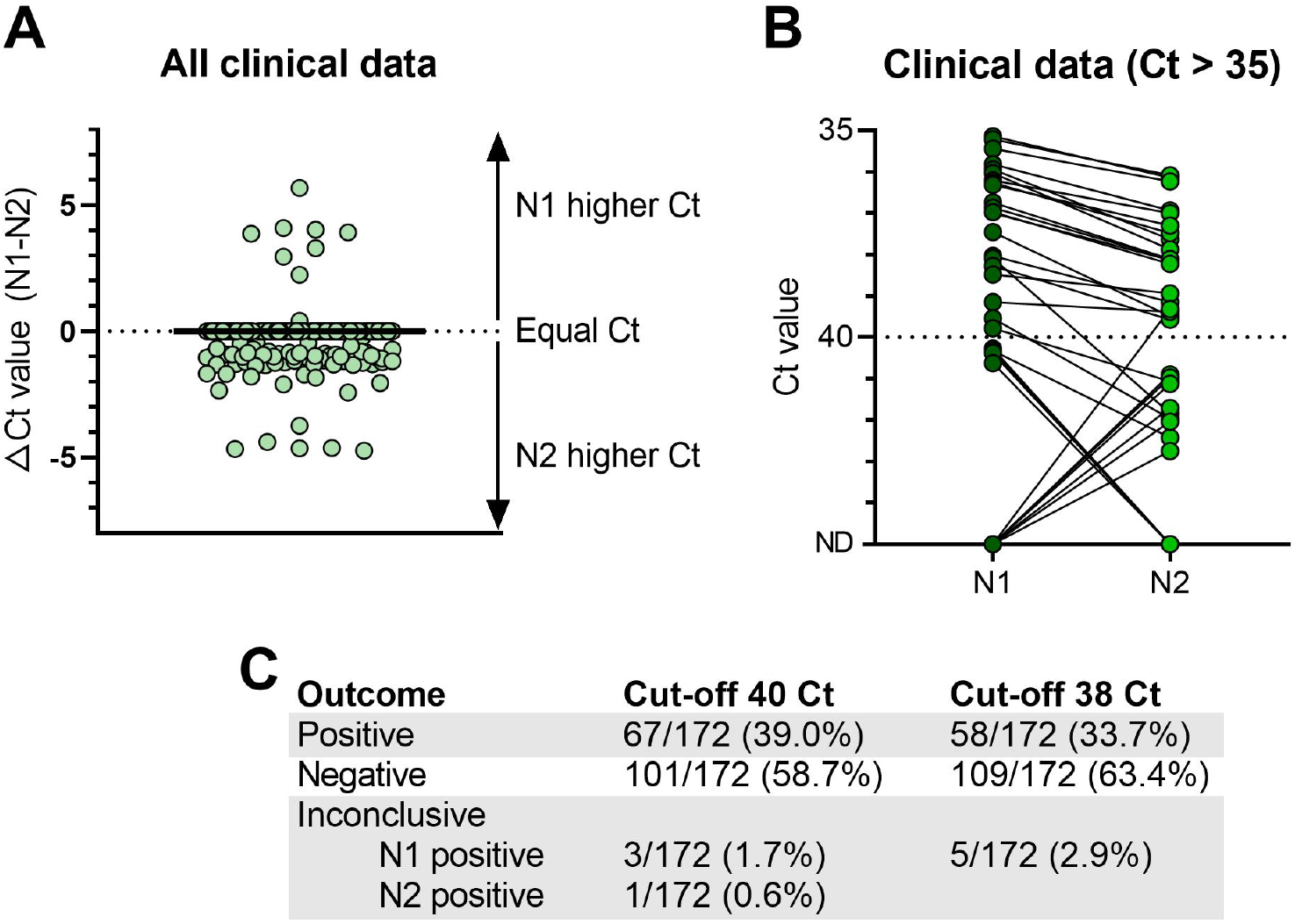
US CDC N1 and N2 primer-probe sets provide consistent test outcomes with clinical samples. We compared Ct values for clinical samples obtained with the 2019-nCoV_N1 (N1) and 2019-nCoV_N2 (N2) primer-probe sets to investigate whether slight differences in analytical sensitivity could lead to inconclusive results. (**A**) We determined the difference in Ct values between N1 and N2 primer probe sets for all tested clinical samples. (**B**) We compared Ct values obtained with the two primer-probe sets for clinical samples with Ct values higher than 35. (**C**) We evaluated outcomes of the US CDC assay based on N1 and N2 at two different cut-off levels (Ct = 40 or 38). We found that N2 has a broader range of Ct values between 40-45, whereas N1 only detected Ct values just above 40. We conclude that these differences do not affect the overall performance of the US CDC assay as the percentage of inconclusive samples is below 3% for cut-off values of 40 or more strictly 38 Ct. N1 = 2019-nCoV_N1, N2 = 2019-nCoV_N2, ND = not detected. Solid black line indicates the median, and dashed line indicates the detection limit.

Using nasopharyngeal swabs, saliva, urine, and rectal swabs from patients as well as nasopharyngeal swabs and saliva from health care workers enrolled in our COVID-19 research protocol at the Yale-New Haven Hospital, we found that more samples had lower Ct values (more efficient virus detection) using the 2019-nCoV_N1 primer-probe set as compared to 2019-nCoV_N2 (**Fig. 4A**). Interestingly, with the N1 set, samples with a Ct value of approximately 40 and samples that were not detected (ND) were clearly discrete groups, whereas several samples that were not detected by the N1 set were in the 41-43 Ct range for the N2 set (**Fig. 4B**). When we look at the US CDC assay outcomes, which take into account both the N1 and N2 results, only 1 out of 172 tests was deemed inconclusive due to N1 being negative (>40 Ct) and N2 being positive (<40 Ct; **Fig. 4C**). We found more inconclusive results where N1 was the only positive set at both 40 Ct (3/172) and 38 Ct (5/172) cut-offs (**Fig. 4C**), likely because the 2019-nCoV_N1 primer-probe set is more sensitive (**Figs. 3, 4A, 4B**). Overall, we generated inconclusive results from less than 3% of the tested clinical samples using the US CDC primer-probe sets, indicating that the US CDC N1 and N2 primer-probe sets are highly consistent for clinical testing.

### Lower sensitivity of RdRp-SARSr (Charité) primer-probe set

To further investigate the relatively low performance of the RdRp-SARSr (Charité) primer-probe set, we compared our standardized primer-probe concentrations with the recommended concentrations in the confirmatory (Probe 1 and Probe 2) and discriminatory (Probe 2 only) RdRp-SARSr (Charité) assays. We deviated from the recommended concentrations in the original assays to make a fair comparison across primer-probe sets, using 500 nM of each primer and 250 nM of probe 2. To investigate the effect of primer-probe concentration on the ability to detect SARS-CoV-2, we made a direct comparison between (***1***) our standardized primer (500 nM) and probe (250 nM) concentrations, (***2***) the recommended concentrations of 600 nM of forward primer, 800 nM of reverse primer, and 100 nM of probe 1 and 2 (confirmatory assay), and (***3***) the recommended concentrations of 600 nM of forward primer, 800 nM of reverse primer, and 200 nM of probe 2 (discriminatory assay) per reaction^5^. We found that adjusting the primer-probe concentrations or using the combination of probes 1 and 2 did not increase SARS-CoV-2 RNA detection when using 10-fold serial dilutions of our RdRp RNA transcripts, or full-length SARS-CoV-2 RNA from cell culture (**Fig. 5**). The Charité Institute of Virology Universitätsmedizin Berlin assay is designed to use the E-Sarbeco primer-probes as an initial screening assay, and the RdRp-SARSr primer-probes as a confirmatory test^5^. Our data suggest that the RdRp-SARSr assay is not a reliable confirmatory assay at low SARS-CoV-2 amounts.

**Fig 5:**
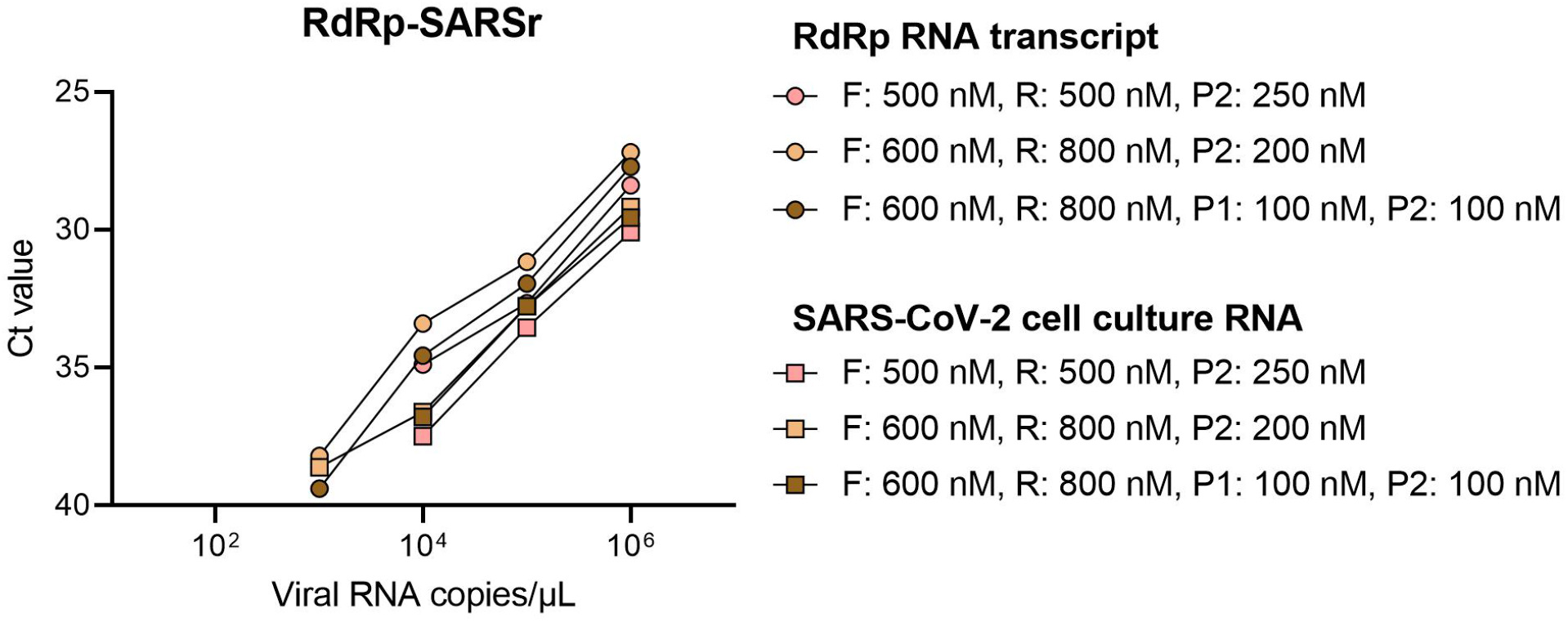
No effect of different concentrations of RdRp-SARSr primers and probes on analytical sensitivity. Low performance of the standardized RdRp-SARSr primer-probe set triggered us to further investigate the effect of primer concentrations. We compared our standardized primer-probe concentrations (500 nM of forward and reverse primers, and 250 nM of probe) with the recommended concentrations in the confirmatory assay (600 nM of forward primer, 800 nM of reverse primer, 100 nM of probe 1, and 100 nM of probe 2), and the discriminatory assay (600 nM of forward primer, 800 nM of reverse primer, and 200 nM of probe 2) as developed by the Charité Institute of Virology Universitätsmedizin Berlin. Standard curves for both RdRp-transcript standard and full-length SARS-CoV-2 RNA are similar, which indicates that higher primer concentrations did not improve the performance of the RdRp-SARSr set. Symbol indicates tested sample type (circles = RdRp transcript standard, and squares = full-length SARS-CoV-2 RNA from cell culture) and colors indicate the different primer and probe concentrations.

### Mismatches in primer binding regions

As viruses evolve during outbreaks, nucleotide substitutions can emerge in primer or probe binding regions that can alter the sensitivity of PCR assays. To investigate whether this had already occurred during the early COVID-19 pandemic, we calculated the accumulated genetic diversity from 992 available SARS-CoV-2 genomes (**Fig. 6A**) and compared that to the primer and probe binding regions (**Fig. 6B**). Thus far we detected 12 primer-probe nucleotide mismatches that have occurred in at least two of the 992 SARS-CoV-2 genomes.

**Fig. 6:**
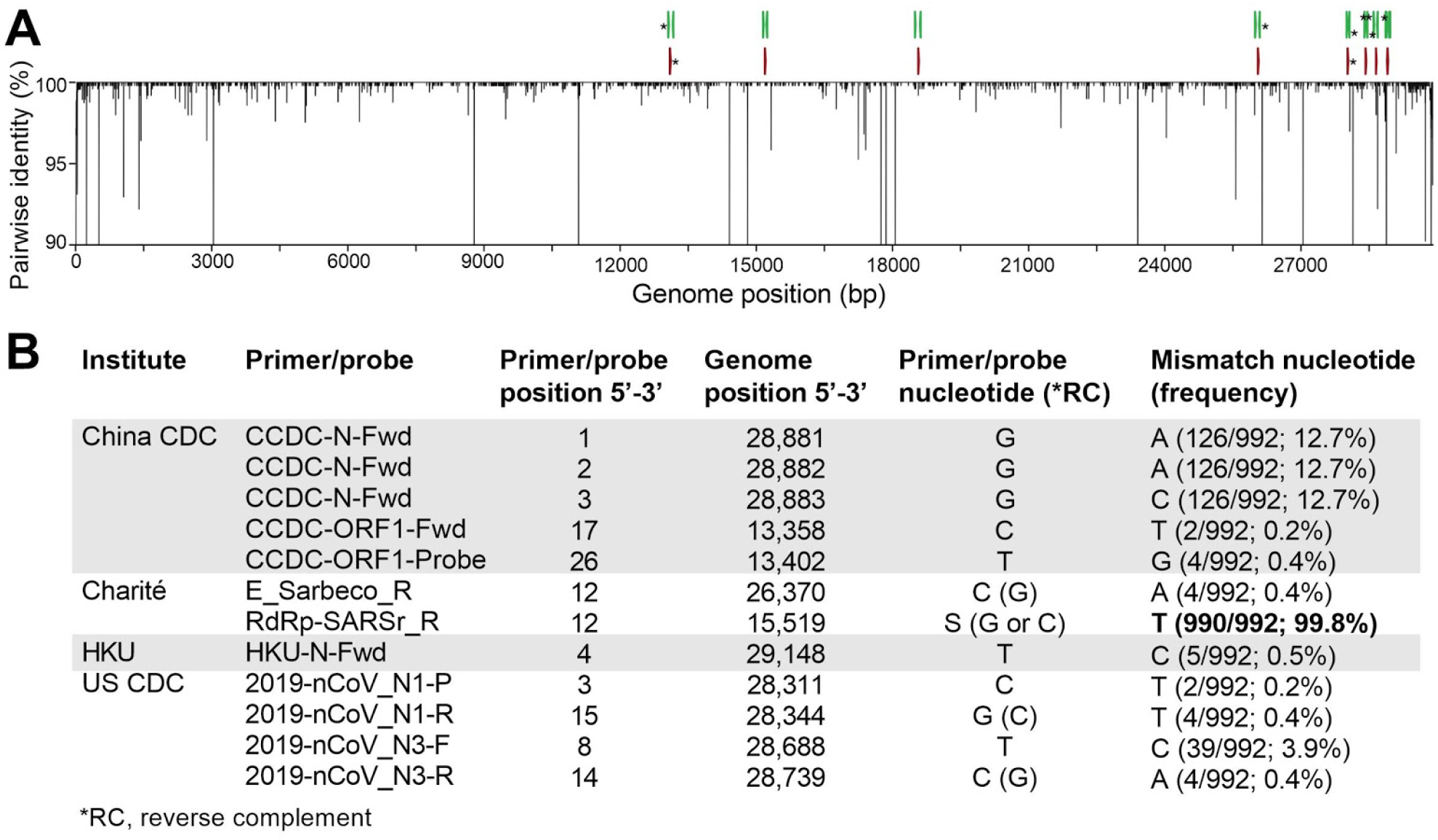
High frequency primer and probe mismatches may result in decreased sensitivity for SARS-CoV-2 detection. (**A**) We aligned nucleotide diversity across 992 SARS-CoV-2 genomes sequenced up to 22 March 2020 and determined mismatches with the nine primer-probe sets. We measured diversity using pairwise identity (%) at each position, disregarding gaps and ambiguous nucleotides. Asterisks (*) at the top indicate primers and probes targeting regions with one or more mismatches. Genomic plots were designed using DNA Features Viewer in Python^16^. (**B**) We only listed mismatch nucleotides with frequencies above 0.1%. These mismatches may result in decreased sensitivity of primer-probe sets.

The most potentially problematic mismatch is in the RdRp-SARSr reverse primer (**Fig. 6B**), which likely explains our sensitivity issues with this set (**Figs. 2, 3, 5**). Oddly, the mismatch is not derived from a new variant that has arisen, but rather that the primer contains a degenerate nucleotide (S, binds with G or C) at position 12, and 990 of the 992 SARS-CoV-2 genomes encode for a T at this genome position (**Fig. 6B**). This degenerate nucleotide appears to have been added to help the primer anneal to SARS-CoV and bat-SARS-related CoV genomes^5^, seemingly to the detriment of consistent SARS-CoV-2 detection. Earlier in the outbreak, before hundreds of SARS-CoV-2 genomes became available, non-SARS-CoV-2 data were used to infer genetic diversity that could be anticipated during the outbreak. As a result, several of the primers contain degenerate nucleotides (**Supplemental Table 3**). For RdRp-SARSr, adjusting the primer (S→A) may resolve its low sensitivity.

Of the variants that we detected in the primer-probe regions, we only found four in more than 30 of the 992 SARS-CoV-2 genomes (>3%, **Fig. 6B**). Most notable was a stretch of three nucleotide substitutions (GGG→AAC) at genome positions 28,881-28,883, which occur in the three first positions of the CCDC-N forward primer binding site. While these substitutions define a large clade that includes ∼13% of the available SARS-CoV-2 genomes and has been detected in numerous countries^14^, their position on the 5’ location of the primer may not be detrimental to sequence annealing and amplification. The other high frequency variant that we detected was T→C substitution at the 8^th^ position of the binding region of the 2019-nCoV_N3 forward primer, a substitution found in 39 genomes (position 28,688). While this primer could be problematic for detecting viruses with this variant, the 2019-nCoV_N3 set has already been removed from the US CDC assay. We found another seven variants in only five or fewer genomes (<0.5%, **Fig. 6B**), and their minor frequency at present does not pose a major concern for viral detection. This scenario may change if those variants increase in frequency: most of them lie in the second half of the primer binding region, and may decrease primer sensitivity^15^. The WA1_USA strain (GenBank: MN985325) that we used for our comparisons did not contain any of these variants.

## Discussion

Our comparative results of primer-probe sets used in qRT-PCR assays indicate a high similarity in the analytical sensitivities for SARS-CoV-2 detection. We found that the most sensitive primer-probe sets are E-Sarbeco (Charité), HKU-ORF1 (HKU), HKU-N (HKU), CCDC-N (China CDC), 2019-nCoV_N1 (US CDC), and 2019-nCoV_N3 (US CDC), which could partially detect SARS-CoV-2 at 1 (25%) and 10 (25-50%) virus copies per μL of RNA. In contrast, the RdRp-SARSr (Charité) primer-probe set had the lowest sensitivity, as also shown by an independent study^13^, likely stemming from a mismatch in the reverse primer. We found slight differences in analytical sensitivity of 2019-nCoV_N1 and N2, but this did not affect the outcomes of the US CDC assay when testing clinical samples from the COVID-19 pandemic. Overall, our findings indicate that all tested qRT-PCR primer-probe sets are reliable for accurate detection of SARS-CoV-2 and yield comparable results.

Our initial preliminary data indicated background cross reactivity when testing clinical samples from pre-COVID-19 respiratory disease patients with the CCDC-N and CCDC-ORF1 (China CDC), and the 2019-nCoV_N2 and N3 primer-probe sets (US CDC). However, when attempting to increase the number of replicates with the same pooled mock sample as well as newly created pools from different nasopharyngeal swabs (as shown in Fig 3) we haven’t been able to detect any background amplification in “no virus” mock samples. Thus, we believe that our preliminary findings of background amplification were an artifact and, therefore, we did not include these data in the current manuscript.

Our study does have several limitations to consider. First, we standardized PCR conditions to make a fair comparison between primer-probes sets used in four common qRT-PCR assays for detection of SARS-CoV-2. By standardizing the concentration of primers and probes, PCR kits, and thermocycler conditions, we deviated from the conditions as recommended by each assay which may have influenced our findings. For instance, we selected an annealing temperature of 55°C which was lower than recommended for the assays developed by Charité (58°C)^5^ and HKU (60°C)^4^, but similar to the assay developed by US CDC (55°C)^6^. No specific PCR conditions were reported for the assay developed by the China CDC^7^. The two assays (Charité and HKU) with higher annealing temperatures had high analytical sensitivity, which suggests that our standardized annealing temperature likely did not have a large effect on our findings. Second, when determining the sensitivity of primer-probe sets, we performed 24 replicates with no spiked-in virus, and eight replicates at low concentrations of SARS-CoV-2 RNA spiked into (pre-COVID-19) clinical samples. While we evaluated the US CDC using 172 clinical samples collected during the COVID-19 pandemic, more replicates for the other assays are required to accurately determine the lower detection limit. Importantly, analytical sensitivity as reported in our study may not be applicable to other PCR kits or thermocyclers; analytical sensitivities and positive-negative cut-off values should be locally validated when establishing these assays.

In the US, we recommend using the US CDC SARS-CoV-2 assay as (*1*) we found similar analytical sensitivity as compared to the other three assays, (*2*) it includes a human RNase P primer-probe set (RP) that allows for quality control of RNA extraction methods, and (*3*) its wide-spread use in the US makes it easier to compare results. In other regions of the world, however, a different test may be preferable based on existing usage.

## Methods

### Ethics

Residual de-identified nasopharyngeal samples collected during 2017 (pre-COVID-19) were obtained from the Yale-New Haven Hospital Clinical Virology Laboratory. In accordance with the guidelines of the Yale Human Investigations Committee (HIC), this work with de-identified samples is considered non-human subjects research. These samples were used to create the mock substrate for the SARS-CoV-2 spike-in experiments (**Figs. 2, 3**). Clinical samples from COVID-19 patients during March 2020 at the Yale-New Haven Hospital were collected in accordance to the HIC-approved protocol #2000027690. These samples were used to test the US CDC 2019-nCoV_N1 and 2019-nCoV_N2 primer-probe sets (**Fig. 4**).

### Generation of RNA transcript standards

We generated RNA transcript standards for each of the five genes targeted by the diagnostic qRT-PCR assays using T7 transcription. A detailed protocol can be found here^10^. Briefly, cDNA was synthesized from full-length SARS-CoV-2 RNA (WA1_USA strain from UTMB; GenBank: MN985325). Using PCR, we amplified the nsp10, RdRp, nsp14, E, and N genes with specifically designed primers (**Supplemental Table 1**). We purified PCR products using the Mag-Bind TotalPure NGS kit (Omega Bio-tek, Norcross, GA, USA) and quantified products using the Qubit High Sensitivity DNA kit (ThermoFisher Scientific, Waltham, MA, USA). We determined fragment sizes using the DNA 1000 kit on the Agilent 2100 Bioanalyzer (Agilent, Santa Clara, CA, USA). After quantification, we transcribed 100-200 ng of each purified PCR product into RNA using the Megascript T7 kit (ThermoFisher Scientific). We quantified RNA transcripts using the Qubit High sensitivity RNA kit (ThermoFisher Scientific) and checked quality using the Bioanalyzer RNA pico 6000 kit. For each of the RNA transcript standards (**Supplemental Table 2**), we calculated the number of viral RNA copies per µL using Avogadro’s number. We generated a genomic annotation plot with all newly generated RNA transcript standards and the nine tested primer-probe sets based on the NC_045512 reference genome using the DNA Features Viewer Python package (**Fig. 1A**)^16^. We generated standard curves for each combination of primer-probe set with its corresponding RNA transcript standard (**Fig. 1B**), using standardized qRT-PCR conditions as described below.

### qRT-PCR conditions

To make a fair comparison between nine primer-probe sets (**Table 1)**, we used the same qRT-PCR reagents and conditions for all comparisons. We used the Luna Universal Probe One-step RT-qPCR kit (New England Biolabs, Ipswich, MA, USA) with 5 µL of RNA and standardized primer and probe concentrations of 500 nM of forward and reverse primer, and 250 nM of probe for all comparisons. PCR cycler conditions were reverse transcription for 10 minutes at 55°C, initial denaturation for 1 min at 95°C, followed by 40 cycles (45 cycles for clinical samples) of 10 seconds at 95°C and 20 seconds at 55°C on the Biorad CFX96 qPCR machine (Biorad, Hercules, CA, USA). We applied the fluorescence drift correction for plates with autofluorescence and refrained from manual adjustment of the threshold. We calculated analytical efficiency of qRT-PCR assays tested with corresponding RNA transcript standards using the following formula^17,18^:

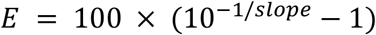

### Validation with SARS-CoV-2 RNA and pre-COVID-19 mock samples

We prepared mock samples by extracting RNA from de-identified nasopharyngeal swabs collected in 2017 (pre-COVID-19) from hospital patients with respiratory disease using the MagMAX Viral/Pathogen Nucleic Acid Isolation kit (ThermoFisher Scientific) following manufacturer’s protocol. We used 300 µL of sample and eluted in 75 µL. We compared analytical efficiency and sensitivity of primer-probe sets by testing 10-fold dilutions (10^6^-10^0^ viral RNA copies/μL) of SARS-CoV-2 RNA as well as the SARS-CoV-2 mock samples spiked with RNA after extraction (eluates pooled from 12 individuals), in duplicate. In addition, we pooled eluates from 4 patients to create 4 independent pools (16 individuals total) and spiked these mock samples with 10-fold dilutions of SARS-CoV-2 RNA (10^0^-10^2^ viral RNA copies/μL) to determine the lower detection limit of each primer-probe set. We tested RNA-spiked mock samples from each of the four independent pools in duplicate (in total 8 reps). Lastly, we tested mock samples (no spiked-in virus) from each pool for six replicates (in total 24 reps per primer-probe set) to test for potential background amplification.

### Clinical samples

Clinical samples from COVID-19 diagnosed patients and health care workers were obtained from the Yale-New Haven Hospital. We extracted nucleic acid from nasopharyngeal swabs, saliva, urine, and rectal swabs using the MagMax Viral/Pathogen Nucleic Acid Isolation kit following manufacturer’s protocol. We used 300 µL of each sample and eluted in 75 µL. We used the Luna Universal Probe One-step RT-qPCR kit with standardized primer and probe concentrations of 500 nM of forward and reverse primer, and 250 nM of probe for the 2019-nCoV_N1, 2019-nCoV_N2, and RP (human control) primer-probe sets to detect SARS-CoV-2 in each sample. PCR cycler conditions were reverse transcription for 10 minutes at 55°C, initial denaturation for 1 min at 95°C, followed by 45 cycles of 10 seconds at 95°C and 20 seconds at 55°C on the Biorad CFX96 qPCR machine (Biorad, Hercules, CA, USA). All figures were made with GraphPad Prism 8.3.0.

### Mismatches in primer binding regions

We investigated mismatches in primer binding regions by calculating pairwise identities (%) for each nucleotide position in binding sites of assay primers and probes. Ignoring gaps and ambiguous bases, we compared all possible pairs of nucleotides in all columns of a multiple sequence alignment including all available SARS-CoV-2 genomes from GISAID (as of 22 March 2020). We assigned a score of 1 for each identical pair of bases, and divided the final score by the total number of valid nucleotide pairs, to finally express pairwise identities as percentages. Pairwise identity of less than 100% indicates mismatches between primers or probes and some SARS-CoV-2 genomes. We calculated mismatch frequencies and reported absolute and relative frequencies for mismatches with frequency higher than 0.1%. The DNA Features Viewer package in Python was used to generate the diversity plot (**Fig. 6**)^16^.

## Data Availability

All data are included in this article and the supplemental files.

## Acknowledgements

We thank K. Plante and the University of Texas Medical Branch World Reference Center for Emerging Viruses for providing SARS-CoV-2 RNA, the Yale COVID-19 Laboratory Working Group for technical support, P. Jack and S. Taylor for discussions, and A. Greninger for feedback on a previous version of this manuscript. We also thank scientists from around the world that openly shared their SARS-CoV-2 genomic data on GISAID. This research was funded by the generous support from the Yale Institute for Global Health and the Yale School of Public Health start-up package provided to NDG. CBFV is supported by NWO Rubicon 019.181EN.004.

## Data availability

All data are included in this article and the supplemental files.

## Supplement

**Supplemental Table 1:**
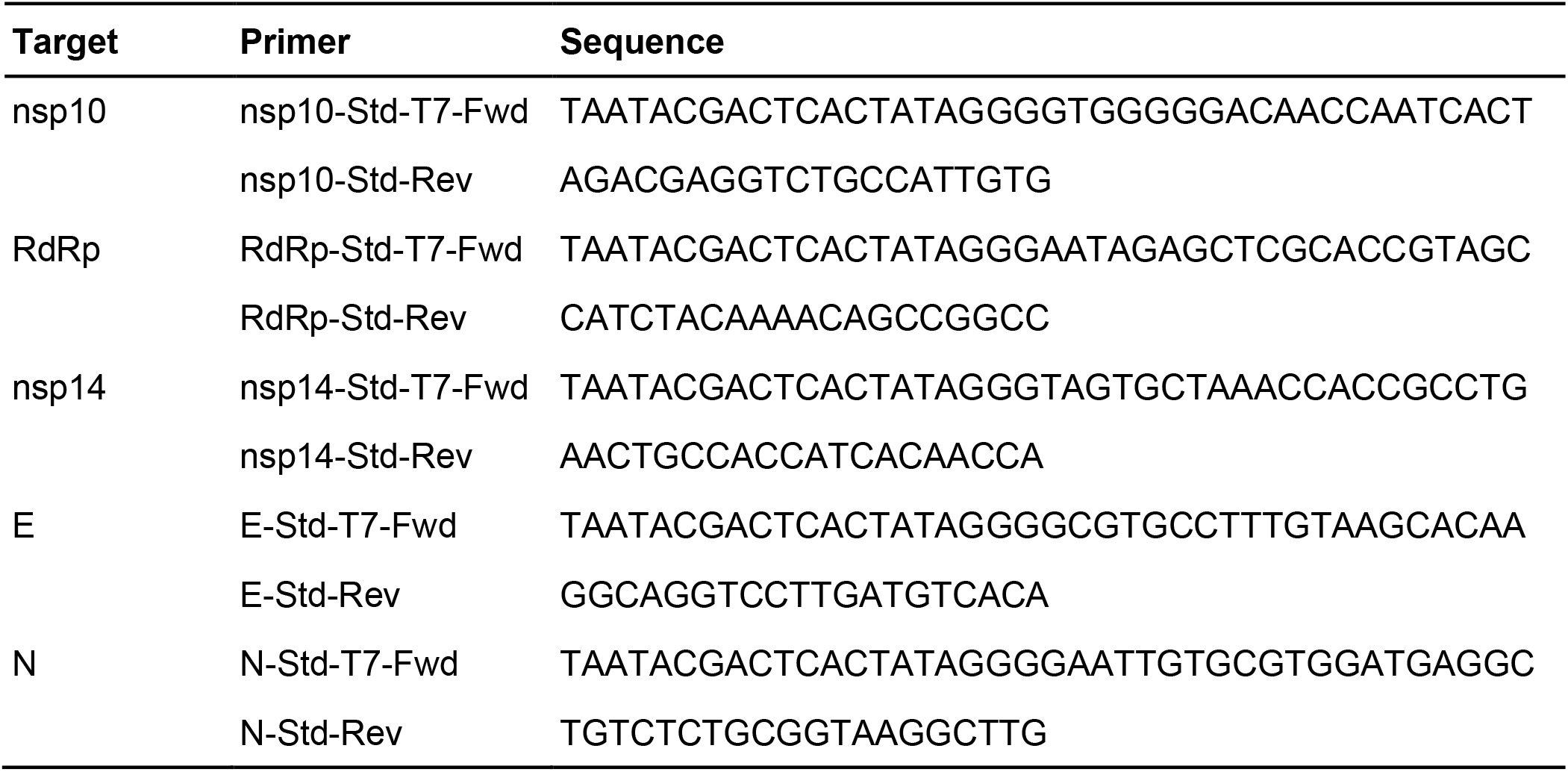
Primers for generation of T7 RNA transcript standards for SARS-CoV-2.

**Supplemental Table 2:**
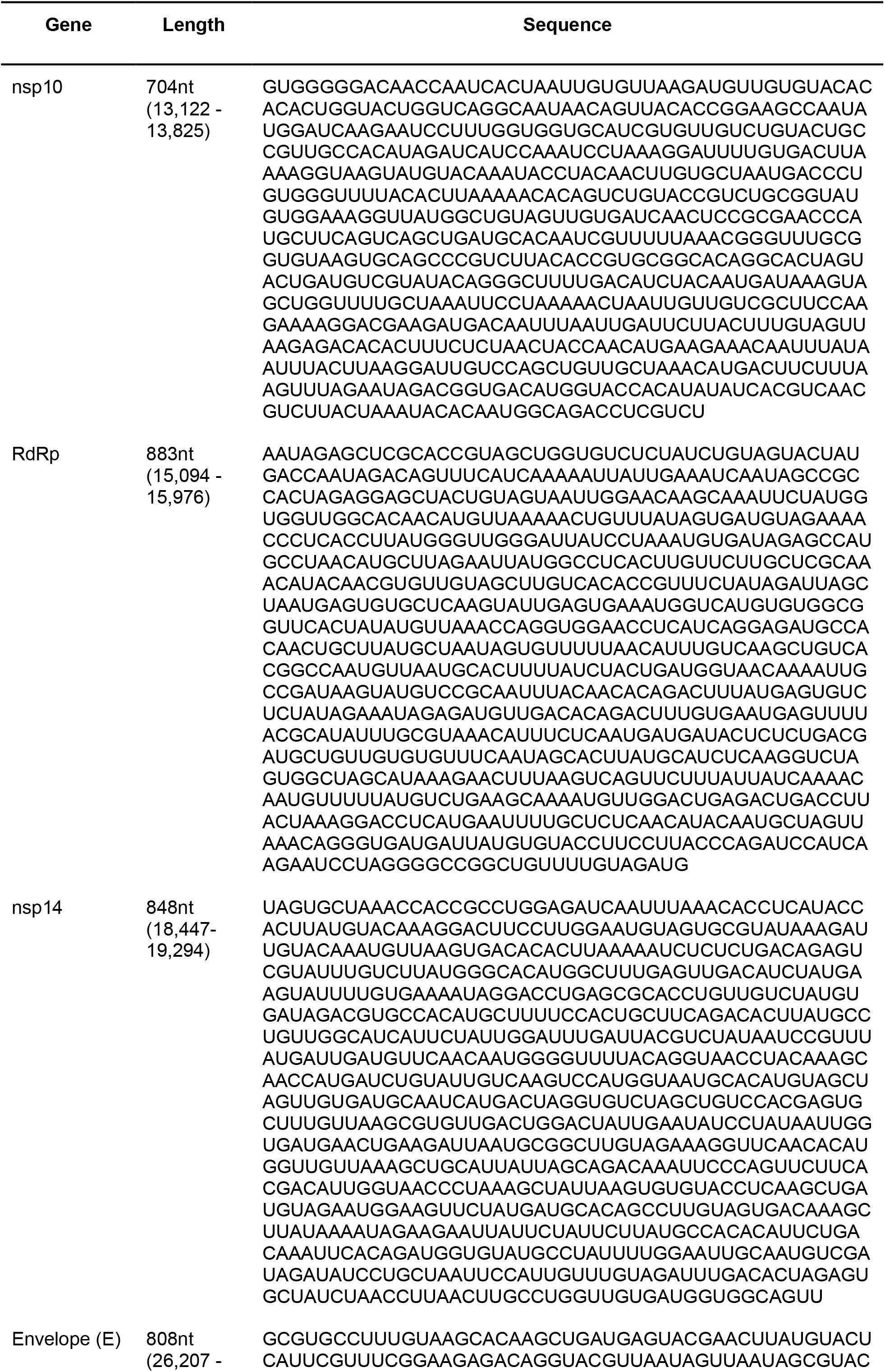
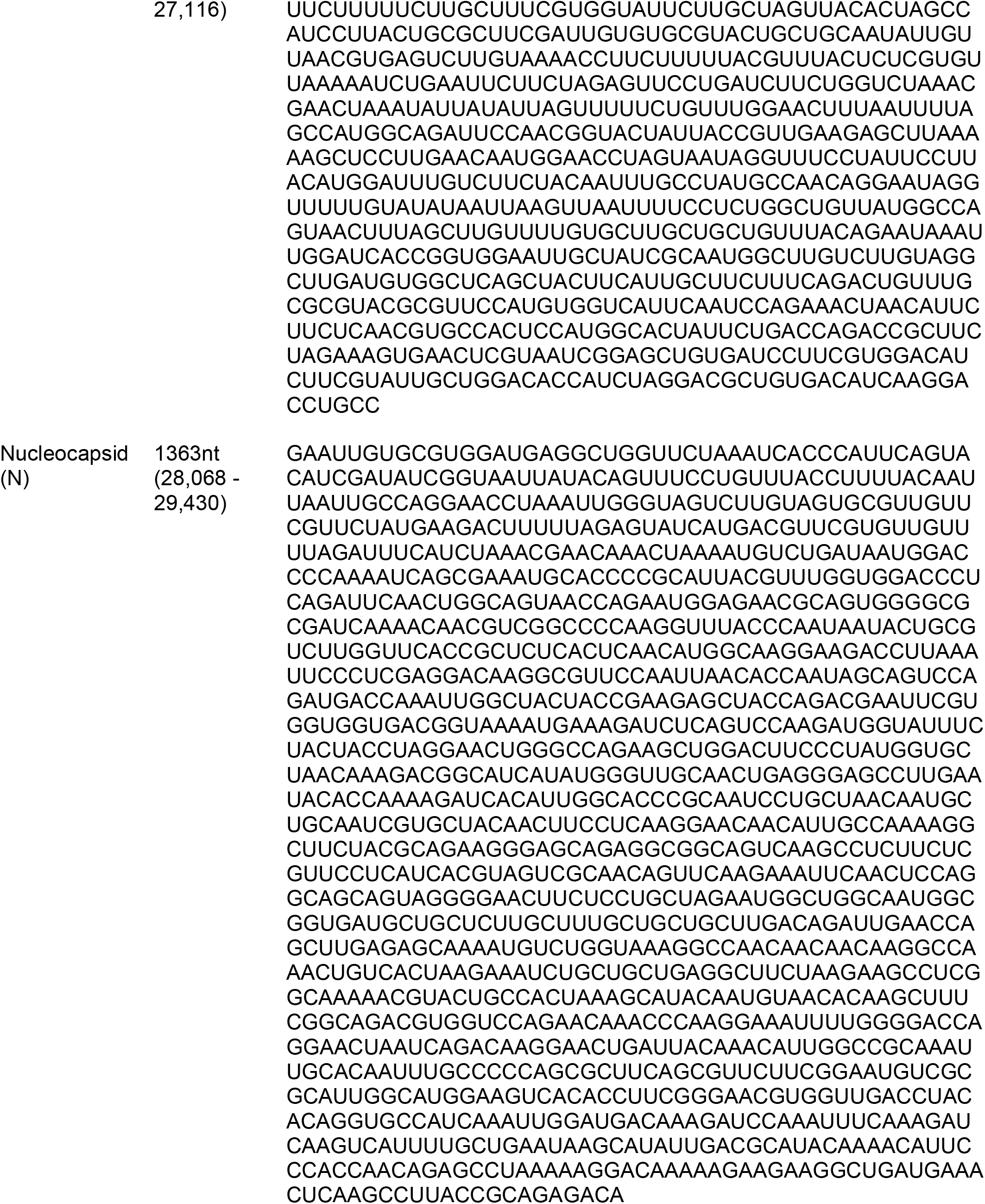
RNA transcript standards for common SARS-CoV-2 diagnostic assays (see genomic context on Figure 1A).

**Supplemental Table 3:** Degenerate bases in common SARS-CoV-2 qRT-PCR assay primers and probes.

| Primer | Degenerate base, and its purpose | Position in primer (5'-3') | Genomic position (5'-3') | Pairing base in genomes (frequency) |
| --- | --- | --- | --- | --- |
| RdRp-SARSr-F | R, to pair with T or C | 5 | 15,435 | T (992/992; 100.0%) |
| RdRp-SARSr-R | **S, to pair with C or G** | 12 | 15,519 | T (990/992; 99.8%) |
| RdRp-SARSr-R | R, to pair with T or C | 3 | 15,528 | T (992/992; 100.0%) |
| HKU-ORF1-F | Y, to pair with A or G | 6 | 18,783 | A (992/992; 100.0%) |
| HKU-ORF1-F | R, to pair with T or C | 12 | 18,789 | T (989/992; 99.7%) |
| HKU-ORF1-P | W, to pair with T or A | 13 | 18,861 | T (992/992; 100.0%) |
| HKU-ORF1-R | R, to pair with T or C | 4 | 18,906 | T (992/992; 100.0%) |
| 2019-nCoV_N3-P | Y, to pair with A or G | 2 | 28,705 | A (992/992; 100.0%) |

